# Modeling Covid-19 incidence by the renewal equation after removal of administrative bias and noise

**DOI:** 10.1101/2022.02.13.22270901

**Authors:** Luis Alvarez, Jean-David Morel, Jean-Michel Morel

## Abstract

The sanitary crisis of the past two years has focused the public’s attention on quantitative indicators of the spread of the COVID-19 pandemic. The daily reproduction number *R*_*t*_, defined by the average number of new infections caused by a single infected individual at time *t*, is one of the best metrics for estimating the epidemic trend. In this paper, we give a complete observation model for sampled epidemiological incidence signals obtained through periodic administrative measurements. The model is governed by the classic renewal equation using an empirical reproduction kernel, and subject to two perturbations: a time-varying gain with a weekly period and a white observation noise. We estimate this noise model and its parameters by extending a variational inversion of the model recovering its main driving variable *R*_*t*_. Using *R*_*t*_, a restored incidence curve, corrected of the weekly and festive day bias, can be deduced through the renewal equation. We verify experimentally on many countries that, once the weekly and festive days bias have been corrected, the difference between the incidence curve and its expected value is well approximated by an exponential distributed white noise multiplied by a power of the magnitude of the restored incidence curve.

**Simple Summary:** In the past two years, the COVID-19 incidence curves and reproduction number *R*_*t*_ have been the main metrics used by policy makers and journalists to monitor the spread of this global pandemic. However, these metrics are not always reliable in the short term, because of a combination of delay in detection, administrative delays and random noise. In this article, we present a complete model of COVID-19 incidence, faithfully reconstructing the incidence curve and reproduction number from the renewal equation of the disease and precisely estimating the biases associated with periodic weekly bias, festive day bias and residual noise.

## 1. Introduction

The renewal equation, first formulated for birth-death processes in a 1907 note of Alfred Lotka [1], establishes a model for epidemic propagation based on the individual infectiousness. The infectiousness of individuals at time *t* is characterized by the reproduction number *R*_*t*_, defined as the average number of cases generated by an infected person at time *t*, and by the generation time, [2,3], defined as the probability distribution of the time between infection of a primary case and infections in secondary cases. This probability distribution depends on the incubation time (a permanent biological factor) and on the detection time (which we assume stationary). For these reasons, the distribution of the generation time is supposed to be independent of *t*. In practice, the generation time is replaced by the observable serial interval Φ_*s*_ which represents the time distribution of the delay of the onset of symptoms between primary and secondary cases. In Fig. 1, we show the serial interval obtained in [4] u0sing 689 observed pairs of primary and secondary cases.

**Figure 1.**
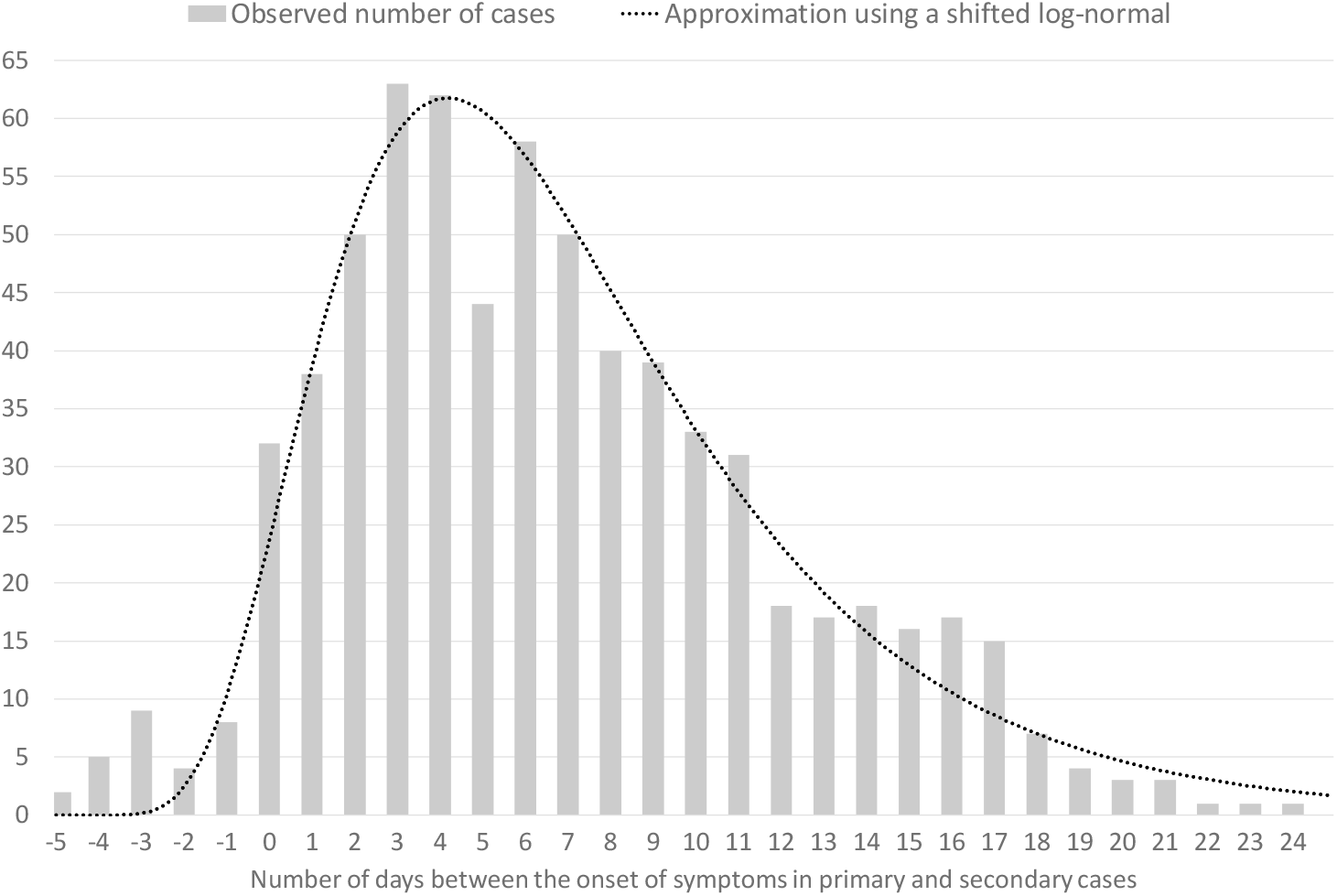
The serial interval Φ_*s*_ obtained by [4]. The bars represent the observed number of cases in function of the number of days between the onset of symptoms in primary and secondary cases. The dotted line is its approximation by a scaled and shifted log-normal distribution.

The case renewal equation [5,6] is a classic equation linking *R*_*t*_, Φ and the incidence *i*_*t*_ of new daily cases,

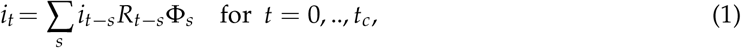

where *t*_*c*_ is the current time. This equation does not account for several strong perturbations of *i*_*t*_. Government statistics of the observed incidence curve are indeed affected by changes in testing and polling policies and by weekend reporting delays. These recording delays and subsequent rash corrections result in impulse noise, and in a strong weekly periodic bias observable on the observed incidence curve 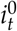. In [3] this bias is corrected by a seven days sliding average and in [7] it is corrected by multiplying 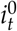 by a 7-day periodic factor *q*_*t*_. These *bias correcting coefficients q*_*t*_ are learned by a variational method that we describe below. Our first purpose in this note is to resolve the festive day problem. We denote by **F** the set of festive days *t*, at which the 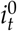 curve is strongly affected by the reduction in the number of registered cases. This reduction is compensated by an increase in the number of registered cases the following days. No model has been proposed so far to address this problem, which creates strong impulse noise in any estimation of *i*_*t*_ and *R*_*t*_. We tackle this problem by a variational method computing *R*_*t*_, where both *i*_*t*_ and *R*_*t*_ are considered unknown on festive days and in the next few days. To that purpose, we shall denote by **F**_+_ the union of festive days and the ones following them affected by the festive day (typically 2 or 3 days after the festive day).

Our second purpose is to give a noise model for the difference 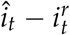 between the signal *î*_*t*_ corrected of the week-end and festive effects, and its restored version 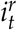 using the renewal equation, defined by

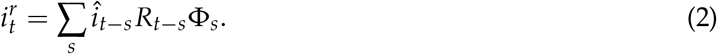

We give strong experimental evidence that the relation between *î*_*t*_ and 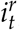, can be empirically modeled by

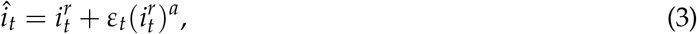

where *a* > 0 and *ε*_*t*_ is a white noise.

This leads us to propose a signal processing version of the renewal equation model taking into account noise and bias and justifying *a posteriori* the variational method. The proposed observation model linking the observed signal 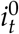 to the ground truth incidence *i*_*t*_ is

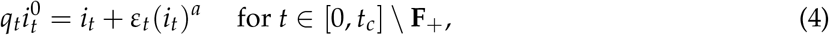

where *q*_*t*_ is a quasi-periodic gain with period 7, *ε*_*t*_ is a white noise. The exponent *a* can be estimated for each country and varies between 0.6 and 0.9. The exceptional set **F**_+_ is introduced because festive days provoke perturbations of the observation model (4). Specifically, the 7 days period of *q*_*t*_ is broken for these groups of days.

We shall verify experimentally on 38 countries (and detail the results on USA, France and Germany) that the normalized error *ε*_*t*_ is indeed a white noise with a distribution that is well described by an exponential distribution. This *a posteriori* noise model contradicts the classic *a priori* stochastic formulation of the renewal equation where the first member *i*_*t*_ of equation (1) is assumed to be a Poisson variable, and the second member of this equation is interpreted as the expectation of this Poisson variable. Using this Poisson model leads to maximum likelihood estimation strategies to compute *R*_*t*_ [3,8–10]. As we shall see, the Poisson model is not verified. Indeed, as we mentioned, the empirically observed standard deviation of the noise follows a power law with exponent *a* significantly larger than 0.5, which is incompatible with the Poisson model.

The proposed observation model (4) of the pandemic’s incidence curve gives a simple framework enabling:

1. a computation of the reproduction number *R*_*t*_;
2. a correction of the weekend and festive days bias on *i*_*t*_;
3. a verification that the difference between the observed incidence curve after bias correction and its expected value using the renewal equation is a white noise, the parameters of which can be estimated.

### Plan

In section 2 we describe an anterior variational method [7] and point out its main three limitations: its weekly bias correction is strongly periodic, which does not work on long periods; the festive days cause strong perturbations in the inversion, finally no residual noise model is proposed. We therefore modify its variational formulation. In section 3 we present the results of the statistical analysis of the residual noise on many countries. These examples lead to specify the noise model and to validate *a posteriori* the proposed inversion model. In section 4 we organize and present all previous *R*_*t*_ and *i*_*t*_ analysis methods, particularly those using an *a priori* noise model. Section 5 is a final discussion.

Timely estimates of restored versions of *i*_*t*_ and *R*_*t*_ are extremely useful to tame a pandemic. The proposed restoration and inversion algorithm can be run through an online demo [11] for every day in every country and U.S. state. The demo plots the objects of this paper, namely the incidence curve 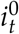, its bias corrected version *î*_*t*_, its fully restored version 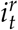, finally the main pandemic index, the time-dependent reproduction number *R*_*t*_. Fig. 2 illustrates the application of the variational method of section 2 to USA on February 1st, 2022, as displayed by the online demo. Fig. 3 compares the results of this inversion method, applied with and without festive day bias correction, obtained for France on January 6, 2022.

**Figure 2.**
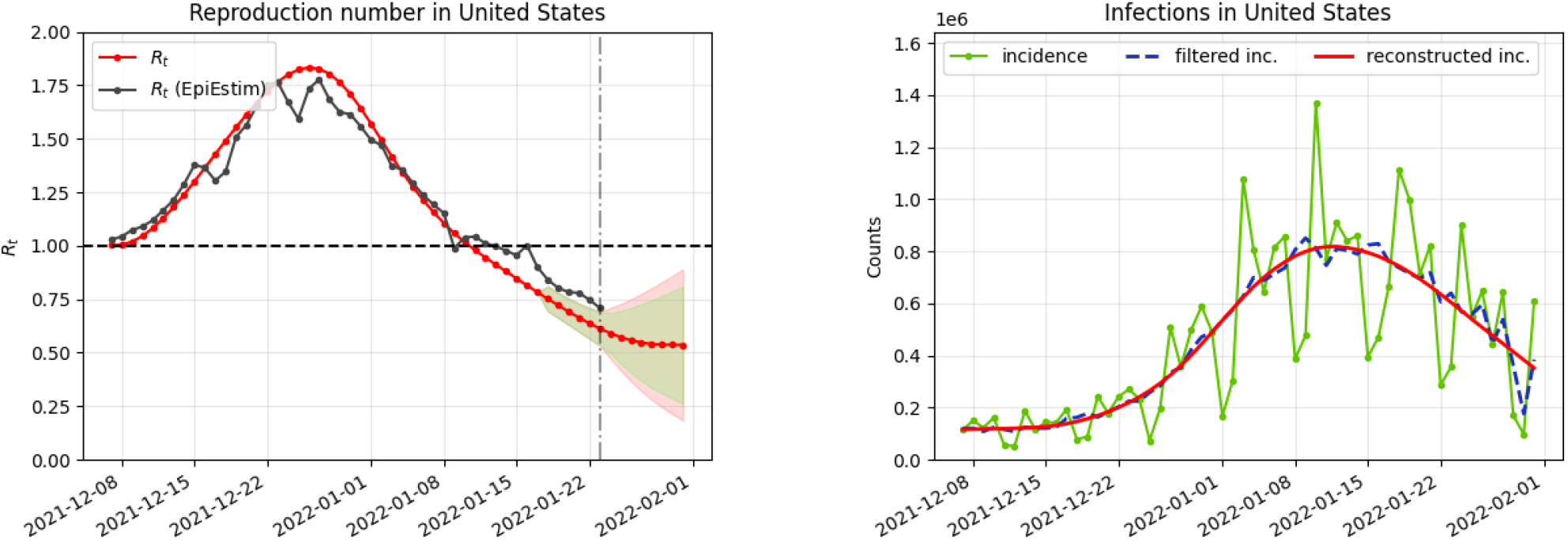
Illustration of the online inversion method [11]. On the left in red, the obtained reproduction number *R*_*t*_ and in black its estimate obtained by the classic EpiEstim method. On the right in green, the original incidence curve *i*_*t*_ of new cases, in blue the incidence curve *î*_*t*_ corrected of the weekend and festive biases, and in red the final reconstructed incidence curve 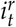 obtained from *R*_*t*_ by the application of the renewal equation. Estimate obtained for USA on February 1st, 2022.

**Figure 3.**
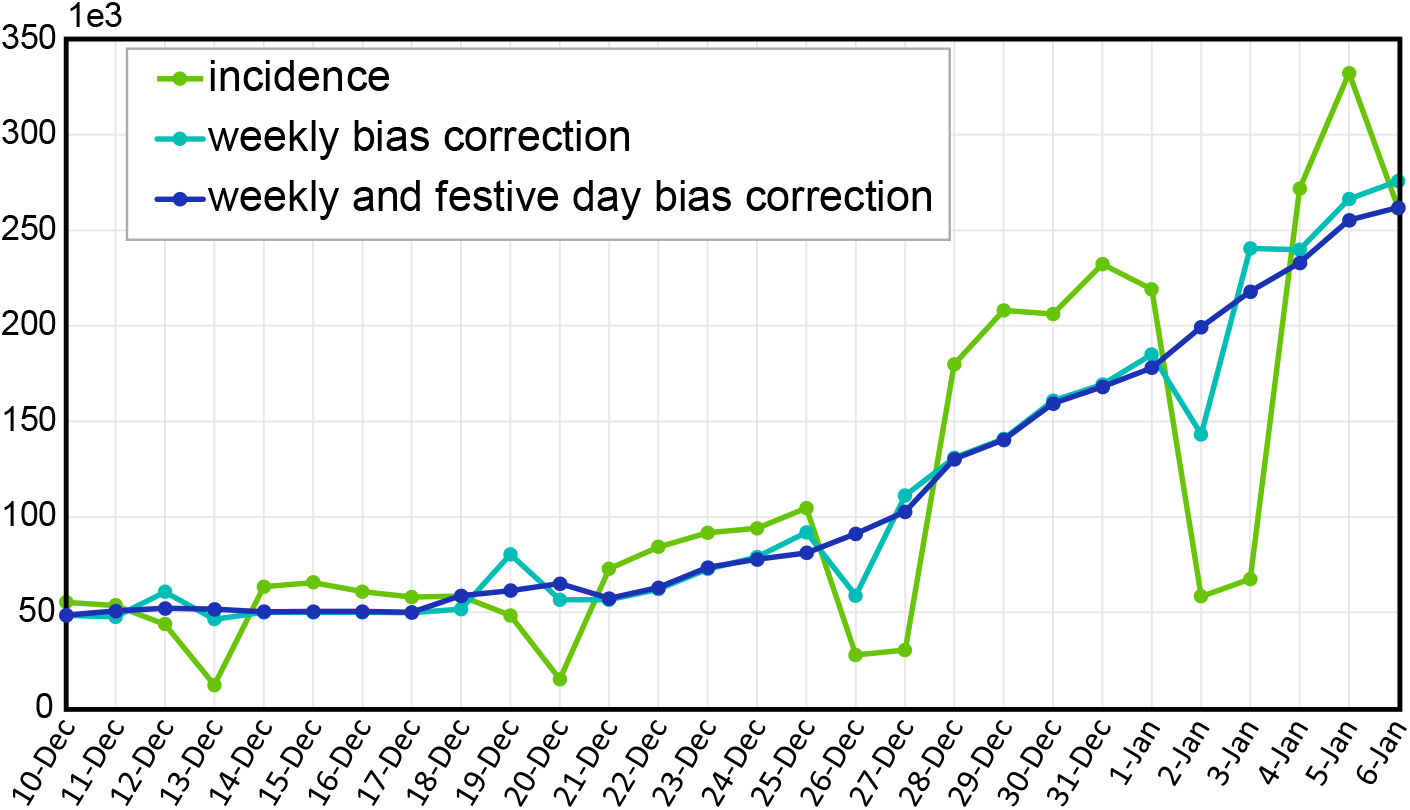
Incidence curve (in green) of France up to January 6, 2022. In cyan, the incidence corrected of the weekly bias, in blue the incidence corrected of the weekly and festive day. The Christmas holidays introduce a distortion in the weekly bias corrected incidence that is corrected by the festive day bias correction.

## 2. The proposed variational model

The EpiInvert method proposed in [7] is a deconvolution + denoising procedure to solve the functional equation (1) using the Tikhonov-Arsenin [12], [13] variational approach. EpiInvert estimates both *R*_*t*_ and a restored *i*_*t*_ corrected for the weekend bias. To remove the weekend effect, it computes a 7-day periodic multiplicative factor *q* = (*q*_0_, *q*_1_, *q*_2_, *q*_3_, *q*_4_, *q*_5_, *q*_6_). From the observed incidence curve and the serial interval, *R*_*t*_ and *q* are jointly estimated by minimizing

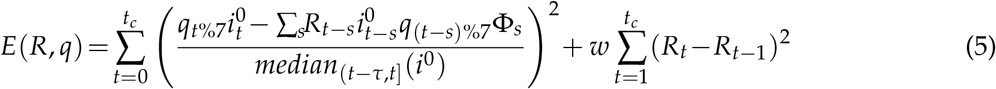

where *t*%7 denotes the remainder of the Euclidean division of *t* by 7 and *median*_(*t*−*τ,t*]_(*i*^0^) is the median of 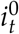 in the interval (*t* − *τ, t*] used to normalize the energy with respect to the size of *i*_*t*_. The total number of cases is preserved by adding to (5) the constraint on *q*_*t*_ :

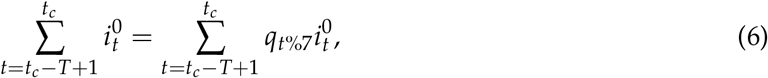

where *T* is a period of analysis empirically fixed to *T* = 56 days. The minimization of the above energy yields estimates of *R*_*t*_, *q* and a restored incidence curve.

One limitation of using a 7-day periodic formulation to model the weekend effect is that it does not take into account the variation over time of the seasonal profile. To deal with this issue, we consider *q*_*t*_ for *t* = 0,.., *t*_*c*_ allowing different correction factors *q*_*t*_ for every day but keeping the values *q*_*t*_ − *q*_*t*−7_ small which forces *q*_*t*_ to be quasi-periodic. A regularity assumption for the seasonality is commonly used in the study of time series as it is the case of the standard Holt-Winters’ seasonal method [14].

In addition to the weekend bias, festive days can introduce a strong bias in the incidence values. On a festive day *t* ∈ **F**, a sharp decrease in the number of registered incident cases is generally observed. This is compensated by increased incidence numbers in the next few days. Assuming that each festive day, *t* ∈ **F**, mainly affects the value of the incidence curve in the festive day and in the next *M*_*t*_ days (where *M*_*t*_ is an algorithm parameter (by default we fix *M*_*t*_ = min{2, *t*_*c*_ − *t*})), we consider the values of 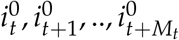 as unknown. We denote by **F**_+_ the union of the festive days *t* ∈ **F** and the *M*_*t*_ days following them. We set 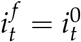 for *t* ∉ **F**_+_ and consider the values 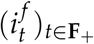 as unknowns. Then the new proposed inversion functional is

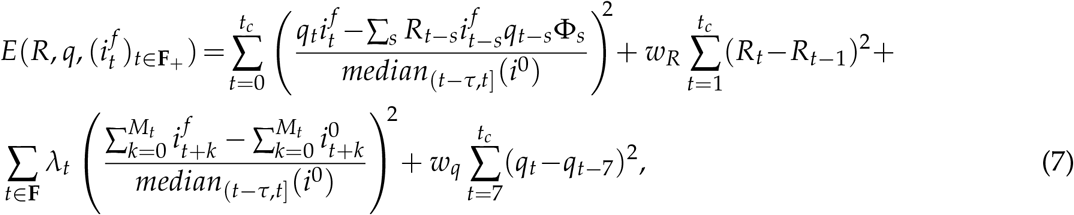

The values 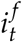 for *t* ∈ **F**_+_ are set free in the minimization. Yet the third term in the functional ensures that the overall number of cases in the affected days remains unchanged. For each *t* ∈ **F**, *λ*_*t*_ ≥ 0 represents the weight we assign to this constraint for each festive day. We fix, experimentally, 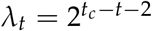 if *t*_*c*_ > *t* and *λ*_*t*_ = 0 if *t*_*c*_ = *t*. In other terms, the value of *λ*_*t*_ is adjusted according to the number of days that have passed since the festive day. To keep a smooth seasonality we add to the energy a regularization term where we penalize high values of *q*_*t*_ − *q*_*t*−7_. The parameters *w*_*R*_ and *w*_*q*_ are regularization weights with default values *w*_*R*_ = *w*_*q*_ = 5. Their values are proven in [7] to be nearly optimal for Covid-19 incidence curves.

By minimizing this energy we obtain the reproduction number *R*_*t*_, the seasonality *q*_*t*_ and 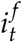, which corresponds to the original incidence 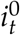 but with the optimized values in the festive days. The bias corrected incidence *î*_*t*_ defined in model (3) is given by 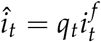.

The estimated incidence curve must preserve the number of cases. In the original EpiInvert formulation this constraint is enforced by (6) on its analysis interval (*t*_*c*_ − *T, T*]. In the new formulation, the interval time of analysis is the whole time interval [0, *t*_*c*_]. Extra conditions are required to keep 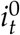 close to *î*_*t*_ and 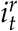. Therefore, to preserve the number of cases we add to the energy (7) the constraints on *q*_*t*_:

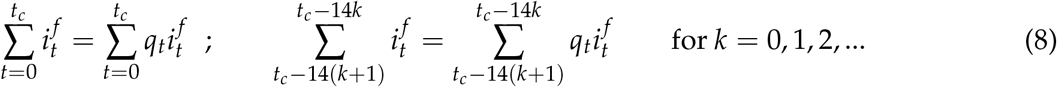

The first constraint corresponds to a global preservation of the number of cases in the whole period and the second one corresponds to a local preservation of the number of cases every 2 weeks. In particular, the second constraint ensures a good agreement between the epidemiological indicator given by the accumulated number of cases in the last 14 days of the original incidence curve and the estimated ones using the proposed method. This indicator is currently widely used to evaluate the current epidemic transmission.

The minimization of the energy (7) is obtained by alternating steps computing in turn *R*_*t*_, *q*_*t*_, and then 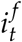 (for *t* ∈ **F**_+_) until convergence. The above constraints are added to the minimization by the Lagrange multiplier technique.

## 3. Results

We used the incidence data published in [15] for France, [16] for Germany, [17] for Spain and [18] for the rest of countries. We checked the observation model and its inversion on the 626 daily incidence data from March 24, 2020 to December 9, 2021 for 38 countries and will detail the results for France, Germany, and the USA. In general, for the festive days we fixed *M*_*t*_ = 2, so the method estimated the incidence value of the festive day and of the next 2 days. However, not all festive days disturb the incidence in the same way. Parameter *M*_*t*_ allows us to adapt the number of days affected. To illustrate this option we set *M*_*t*_ = 5 for Thanksgiving in the USA in 2021 because this festive day causes in 2021 a longer perturbation in the number of registered cases. Figs. 5,A1,A2 show the minimization results for the energy (7). They display for each country (i) the original incidence curve 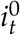, (ii) the incidence curve after bias correction *î*_*t*_, (iii) the restored incidence curve 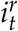 using the renewal equation (1), (iv) the weekly bias correction factors *q*_*t*_, (v) the reproduction number estimation *R*_*t*_ and (vi) the normalized error defined by

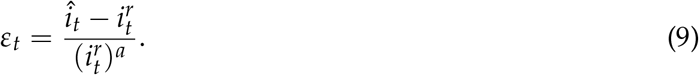

The power *a* was obtained through log-log linear regression. Indeed, if 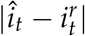 is proportional to 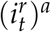, then 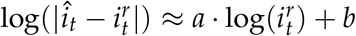, and *a* and *b* can be estimated by a linear regression between 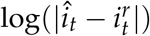 and 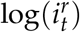. Its results are illustrated for 38 countries in fig. 4 and table A2. The Pearson correlation p-values in this table confirm the linear relation. The estimated exponent *a* varies between 0.7 and 0.9, and the constant coefficient *b* varies between −0.11 and −2.6. For the world we have *a* = 0.76 and *b* = −1.16.

**Figure 4.**
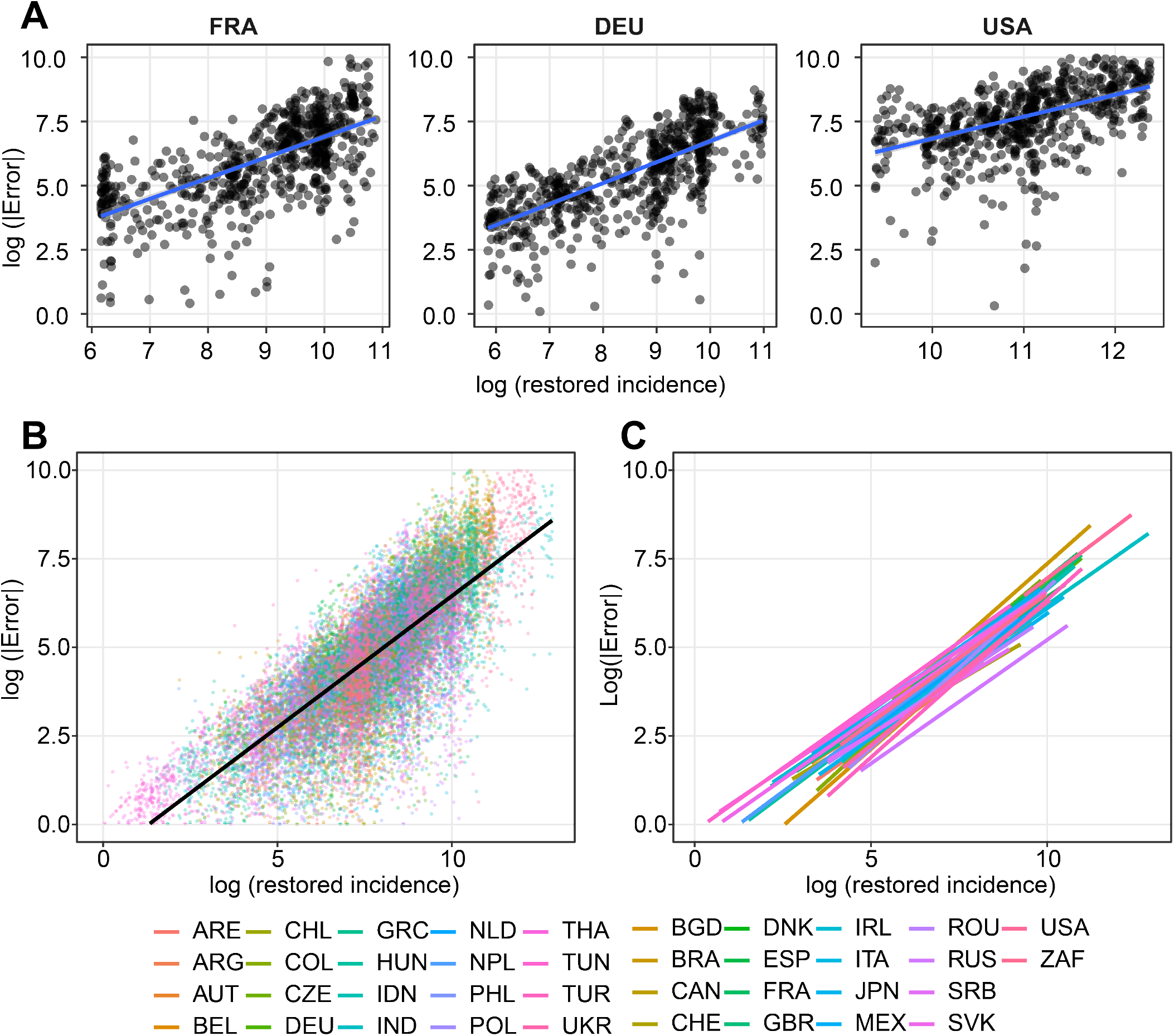
Worldwide log-log correlations between restored incidence 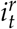 and the residual 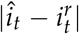 (defined as restored incidence - bias-corrected incidence). The plot presents the log(error) as a function of the log(incidence). The regression parameters were computed through robust linear regression by the R package MASS. A: Correlation in France, Germany, and USA, with festive day correction. B: Spread of the values for 38 countries, without festive corrections. C: Robust linear regression curves for all countries. The linear regression coefficients *a* and *b* can be found in table A2. The worldwide coefficients are *a* = 0.76 and *b* = −1.16.

We performed a control test on a Brownian motion simulated by starting from 10000 and sampling *i*_*t*+1_ − *i*_*t*_ ≃ 𝒩(0, 100). The obtained exponent *a* is negative (*a* = −1.01) and we have *b* = 13.4. Both values are far away from the group of coefficients of real incidence curves. The p-value for the control is anyway non significant (0.0844), compared to the extremely small p-values for the real incidence curves. Fig. A3 shows the results of the variational inversion method on the Brownian control. For this control, both *R*_*t*_ and the weekly seasonality correction coefficients stay very close to 1 as should be expected, with means 1.001 and 1.00002, and standard deviations 1.7% and 0.3% respectively.

Next, we looked for a stochastic model of the normalized error *ε*_*t*_ defined by (9). Figs. 5-A2 visually support a stationarity assumption for *ε*_*t*_ in France, Germany and USA. In fig. 6 we show the autocorrelation function for these three countries. For most non-zero shifts, its value stays inside the 95% confidence interval for the stationarity assumption. (This interval is indicated by horizontal blue lines in the plot.) Similar results were obtained on 33 more countries, as illustrated in fig. A5. These results support a white noise assumption for *ε*_*t*_.

**Figure 5.**
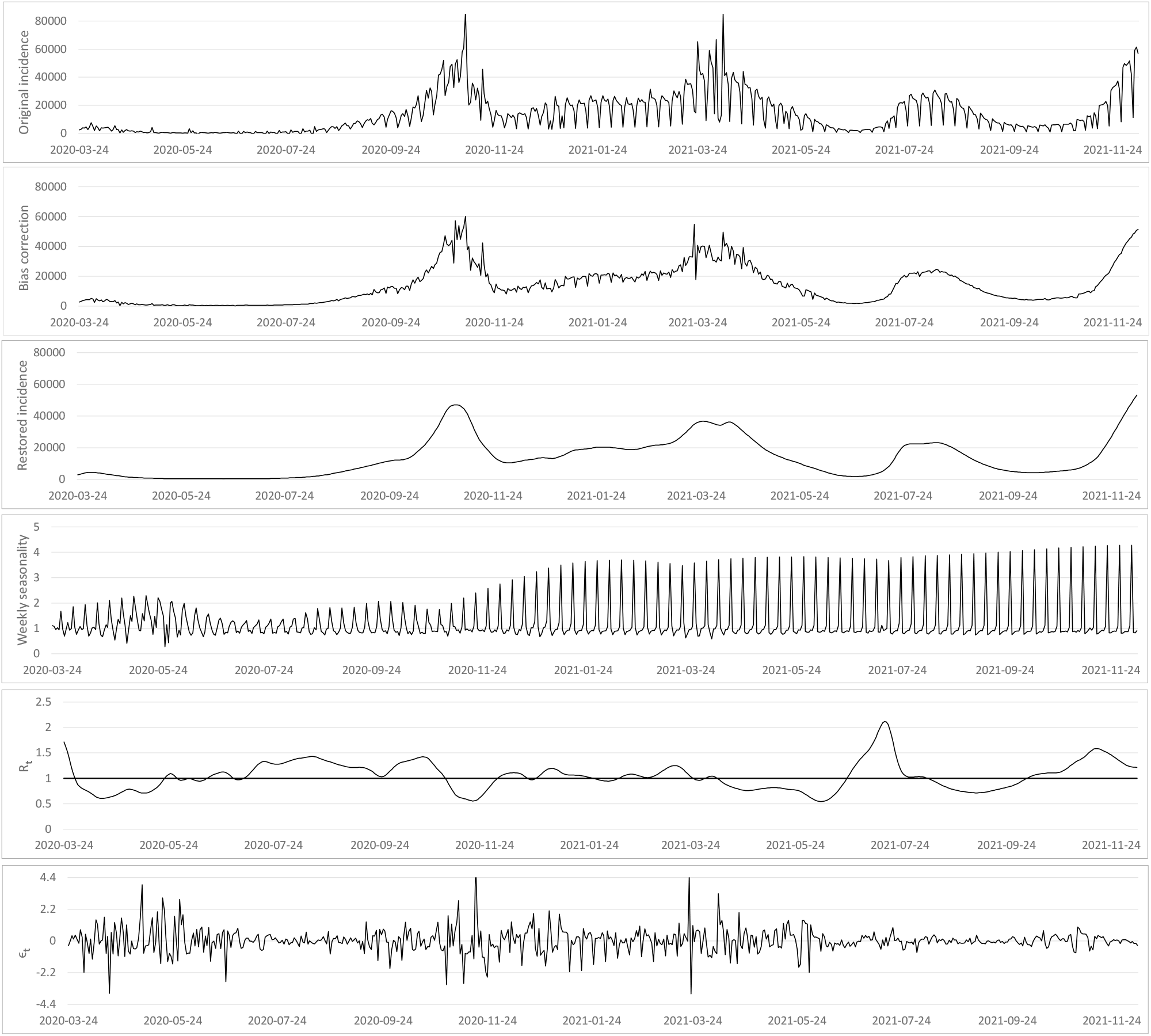
From top to bottom : (i) the original incidence curve 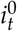 of France, (ii) the incidence curve after bias correction *î*_*t*_, (iii) the restored incidence curve using the renewal equation 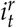, (iv) the weekly bias correction factors *q*_*t*_, (v) the reproduction number estimation *R*_*t*_ and (vi) the normalized error 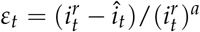, where *a* is the optimal exponent obtained by regression (see table A2).

**Figure 6.**
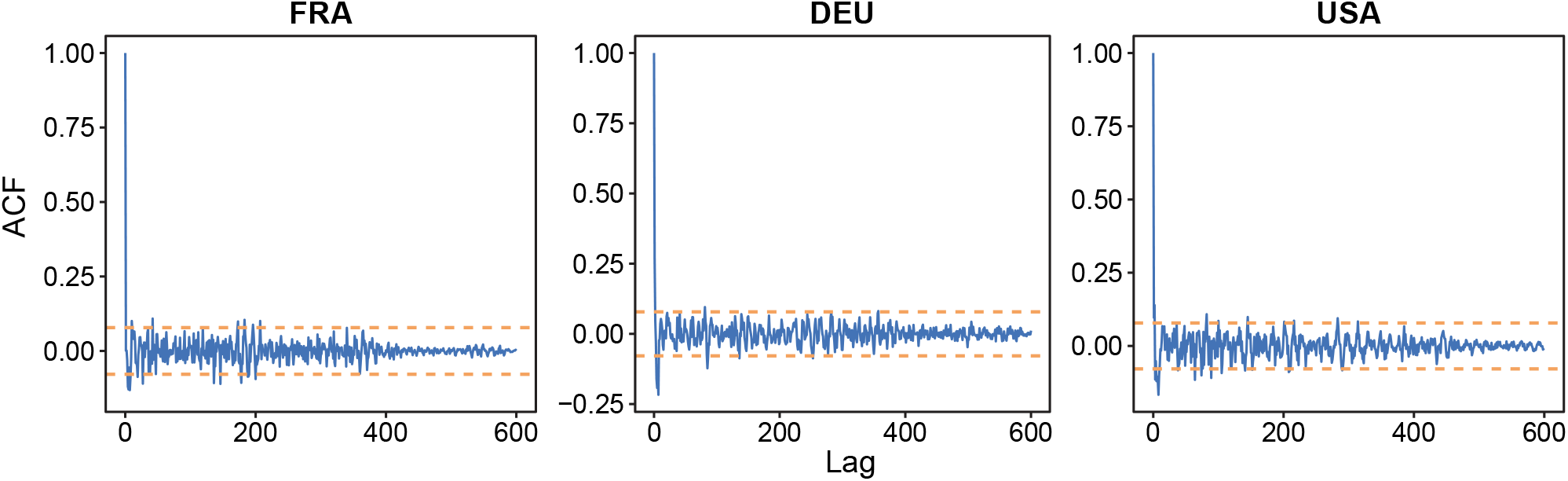
For France, Germany and USA, autocorrelation of the normalized error *ε*_*t*_, using the festive day correction, obtained with the R-software functionalities (acf() function). The orange dotted line gives the 95% confidence interval for non-correlation. Similar plots for the same countries and 33 more countries, without using the festive day correction, are displayed in fig. A5.

We finally estimated the parameters of the distribution of *ε*_*t*_ assuming an exponential power distribution with density

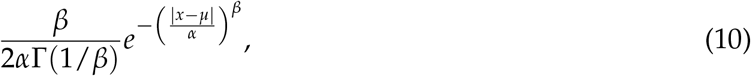

where *μ* is the location, *α* the scale and *β* the shape. These parameters to approximate *ε*_*t*_ by an exponential power distribution were estimated by the R-package *normalp* [19].

In Fig. 7, we plot for these three countries the histogram of the distribution of *ε*_*t*_ and its approximation by a normal (*β* = 2) and by the obtained optimal exponential distribution. We display the same result for 33 more countries in fig. A4.

**Figure 7.**
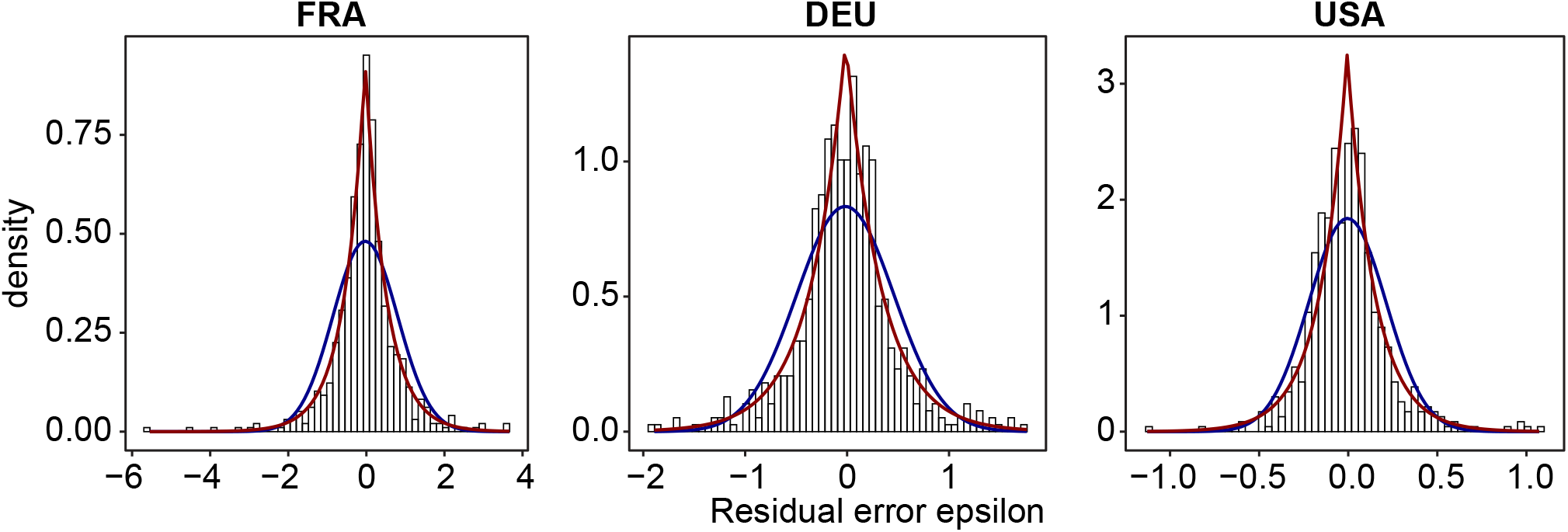
For France, Germany and USA, histogram of the normalized error *ε*_*t*_, using the festive day correction, its normal approximation (blue line) and its optimal approximation using an exponential distribution (red line) (we use the R-package *normalp* to approximate *ε*_*t*_ by an exponential distribution). See fig. A4 for the results for the same countries and 33 more countries without using the festive day correction.

Table A1 gives the results for all countries. Columns 5 to 8 in the table give the parameters of the optimal exponential law: location, scale, shape. In all cases the exponent remains close to 1. Fig. 8 displays a quantile-quantile plot comparing *ε*_*t*_ with the estimated exponential distribution for three countries: France, Germany, USA. The linear fit is excellent, and this goodness of fit is confirmed for 33 more countries in Figure A6.

**Figure 8.**
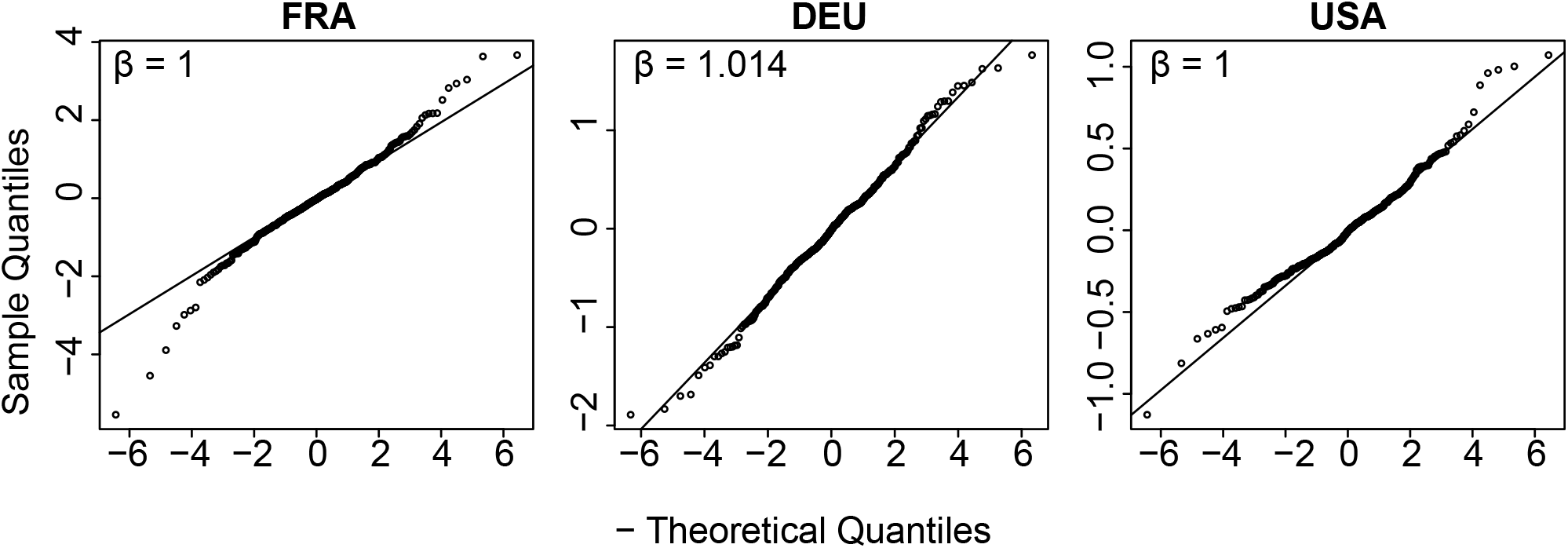
Quantile-quantile plot with France, Germany and the USA comparing *ε*_*t*_, using the festive day correction, with the optimal exponential distribution using the R-package *normalp*.

## 4. Review of previous models

### 4.1. The Fraser renewal equation

In our proposed incidence model, we used the general integral equation (1), which is a functional equation in *R*_*t*_. Integral equations have been previously used to estimate *R*_*t*_: in [20], the authors estimate *R*_*t*_ as the direct deconvolution of a simplified integral equation where *i*_*t*_ is expressed in terms of *R*_*t*_ and *i*_*t*_ in the past, without using the serial interval. A simpler functional equation than (1) was proposed in Fraser [21] (equation (9)),

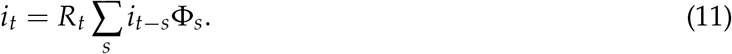

This equation is derived from the general case renewal equation (1) by assuming that *R*_*t*_ is constant in the serial interval support. It computes the “instantaneous reproduction number” and represents the number of secondary cases arising from an individual showing symptoms at a particular time, assuming that conditions remain identical after that time, in contrast with the case renewal equation (1). This last equation applied to the incidence curve is coherent if Φ_*s*_ denotes the serial interval between two cases, which can have negative dates, because an infectious may be detected after the infection cases she caused. Using (11) requires that Φ_*s*_ only has positive dates. This explains why [22] proposed to estimate the generation time, namely the (always positive) time between two infections, before using it in (11). The advantage of equation (11) is that *R*_*t*_ is estimated at time *t* from the past incidence values *i*_*t*−*s*_ by a simple division, provided that Φ_*s*_ = 0 for *s <* 0:

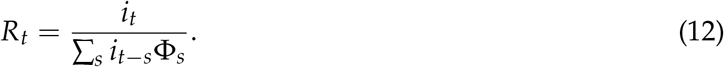

### 4.2. Deterministic implementations using Fraser’s renewal equation (11) and other models

Many papers estimating *R*_*t*_ use the deterministic causal renewal equation (11). This is the case of [23], [24] [25]. This last paper also involves the Wallinga-Teunis formulation [2], also based on the renewal equation but only allowing a backward estimate of *R*_*t*_ (see the discussion in [7]). Some papers like [26] propose a simplified version of (11). See also [27], who use this equation but estimate the probability distribution Φ_*s*_ by a maximum entropy method. A few papers use another deterministic model, the Wallinga-Teunis formulation, to compute *R*_*t*_ [28], or a SIR model, like in [29], where the time variable parameter *β*(*t*) of the three ODE’s of a SIR model is estimated from incidence data in a seven days sliding window.

### 4.3. Stochastic observation models for i_t_ and R_t_

The renewal equation (11) is often endowed with an *a priori* stochastic Poisson model as

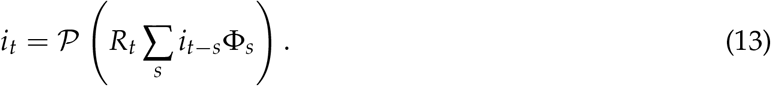

In this stochastic formulation, the first member *i*_*t*_ of Equation (11) is assumed to be a Poisson variable, and the second member of this equation is interpreted as the expectation of this Poisson variable. This leads to a maximum likelihood estimation strategy to compute *R*_*t*_ (see [3,8–10,30]). This form of the renewal equation is proposed and used in [3] and in the EpiEstim software. It is highly recommended in a recent review [31] signed by representatives from ten different epidemiological labs from several continents. Many papers dedicated to the computation of *R*_*t*_ use this model, for example [32], [33] and [34], who also assume that *R*_*t*_ is a Poisson variable, and [35] who also assume that *R*_*t*_ also is a random variable following a Gamma distribution. In [36], the authors use the stochastic form of the renewal equation (13) where they call Φ_*s*_ *causal serial interval*. Then *R*_*t*_ is estimated jointly on all regions of a country by a variational model containing a spatial total variation regularization to ensure that *R*_*t*_ is piecewise constant, and the *L*^1^ norm of its time Laplacian to ensure time regularity. The functional also penalizes outliers, typically Sundays and holidays by assuming a sparse structure of such events. See also [37] for an exposition of the application of this method.

In [38] the method Epifilter is introduced as an extension of EpiEstim and of the Wallinga-Teunis formulation. Epifilter has been applied in practical studies like [39]. The core of Epifilter is again the causal renewal equation in Poisson form (13). Yet, the author proposes a doubly stochastic model, as *R*_*t*_ is assumed to follow a recursive discrete Brownian motion of the sort

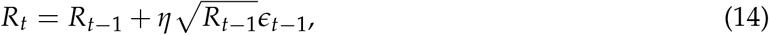

where *ϵ* ≃ 𝒩(0, 1) and *η* is a user parameter, that we can interpret as a regularity control on *R*_*t*_. Then (*R*_*s*_)_*s*≤*t*_ is computed from the incidence data (*i*_*t*_)_*s*≤*t*_ by recursive filtering. The method is complemented by Bayesian (backward) recursive smoothing that brings a better estimate on low incidence periods.

Similarly in [40], a parametric model with a stochastic multiplicative term is proposed for *R*_*t*_ where the stochastic term is a Gamma law with prescribed standard deviation. The parameters are estimated in several prefectures in interaction to give the best fit to incidence data linked to *R*_*t*_ through the causal renewal equation (11).

A few papers assume a negative binomial *a priori* for the incidence [41]. Nevertheless the equations given in the paper indicate the adoption of the renewal equation (11) and put the stochastic process on *R*_*t*_ by assuming *R*_*t*_ ≃ *R*_*t*−1_*GP* where *GP* is a squared exponential kernel. The very same model is used in [42], and is based on the authors’ software EpiNow2. Similarly in [43], incidence *i*_*t*_ and reproduction number *R*_*t*_ are linked through the classic SIR model; a parametric piecewise linear model for *R*_*t*_ is estimated by fitting the parameters to real incidence data. Here the daily incidence data are modeled as a negative binomial, with mean given by the deterministic solution of the SIR equations and unknown dispersion.

In [44], a direct stochastic model is proposed for *R*_*t*_, assuming that its log derivative is Brownian, namely

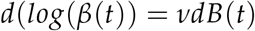

where *ν* is the volatility of the Brownian process to be estimated. Then we have

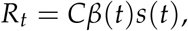

where *C* is a constant depending on steady transmission characteristics and *s*(*t*) is the proportion of the population that is susceptible. The case incidence is then estimated through an SEIR model. We refer to [45] for a still more complex stochastic model for *R*_*t*_, depending on three stochastic parameters.

## 5. Discussion

In [7] we have proven extensively by simulations and experiments on live worldwide Covid-19 incidence data that using the simplified causal renewal equation (11) incurs in a five days delay in the estimation of *R*_*t*_, compared to the Nishiura renewal equation (1). This is why we used here this second model.

All of the stochastic models mentioned in section 4.3 are formulated *a priori*. To the best of our knowledge, no there has been no *a posteriori* verification of their noise models on *i*_*t*_ or *R*_*t*_. In contrast, we have proposed to learn the noise model from data and to verify *a posteriori* that the noise model is correct. Our experiments show that the weekly and festive administrative perturbations are more important than the noise. Hence they must be corrected first to enable a proper noise analysis.

These experiments seem to confirm the validity of the observation model (4). As we saw, this model can be inverted by minimizing the energy (7). This minimization yields three signals: a restored incidence on the festive days, the administrative bias correcting coefficients *q*_*t*_ that are quasi-periodic with period 7, and the time varying reproduction number *R*_*t*_, arguably the pandemic’s most useful control parameter. Last but not least, the renewal equation deduces a restored incidence 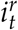 by (2) from the bias compensated incidence *î*_*t*_. The modeling loop was closed by verifying that the normalized error defined by (9) is a white noise. We also found that this noise follows an exponential distribution. This analysis discards the Poisson model for the pandemic’s case count *i*_*t*_. A pure case count *should* be a Poisson noise, but we saw that the main perturbation was an administrative bias which, once compensated, leaves behind a noise with standard deviation proportional to a power larger than 0.5 of the case count *i*_*t*_. Under the Poisson model this standard deviation would have been equal to the square root of *i*_*t*_.

In summary, based on the renewal equation inversion, this work contributes to a better understanding of the dynamic of the registered administrative observation of the incidence curve, its weekly seasonality, the influence of the festive days and the expected noise model in the observation of the incidence curve.

## Data Availability

We use COVID-19 incidence data avaliable at "Our World in Data" webpage.

https://github.com/owid/covid-19-data/tree/master/public/data

---

**Figure A1.**
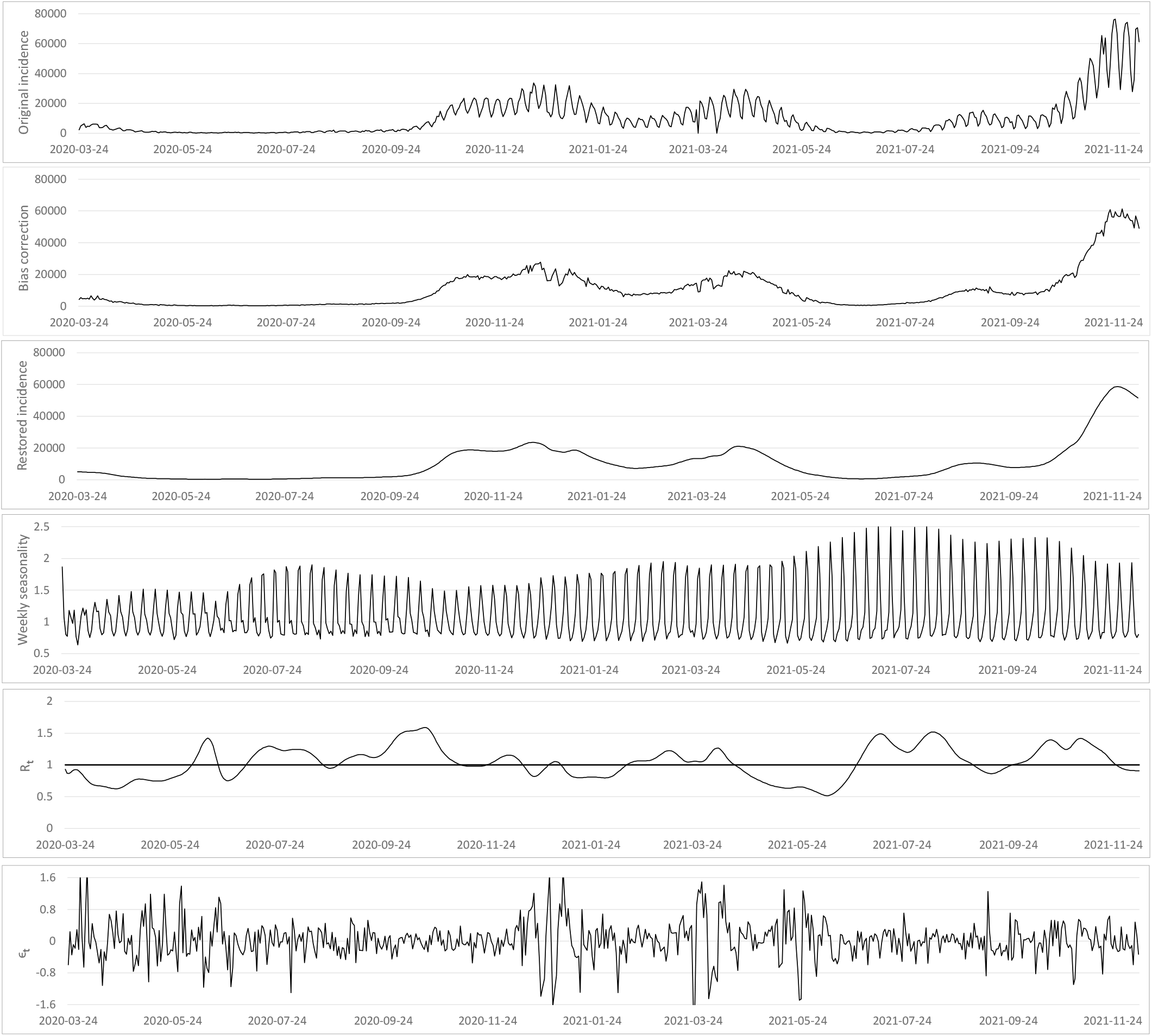
From top to bottom : (i) the original incidence curve of Germany 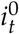, (ii) the incidence curve after bias correction *î*_*t*_, (iii) the restored incidence curve using the renewal equation 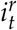, (iv) the weekly bias correction factors *q*_*t*_, (v) the reproduction number estimation *R*_*t*_ and (vi) the normalized error 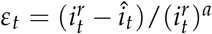, where *a* is the optimal exponent obtained by regression (see table A2).

**Figure A2.**
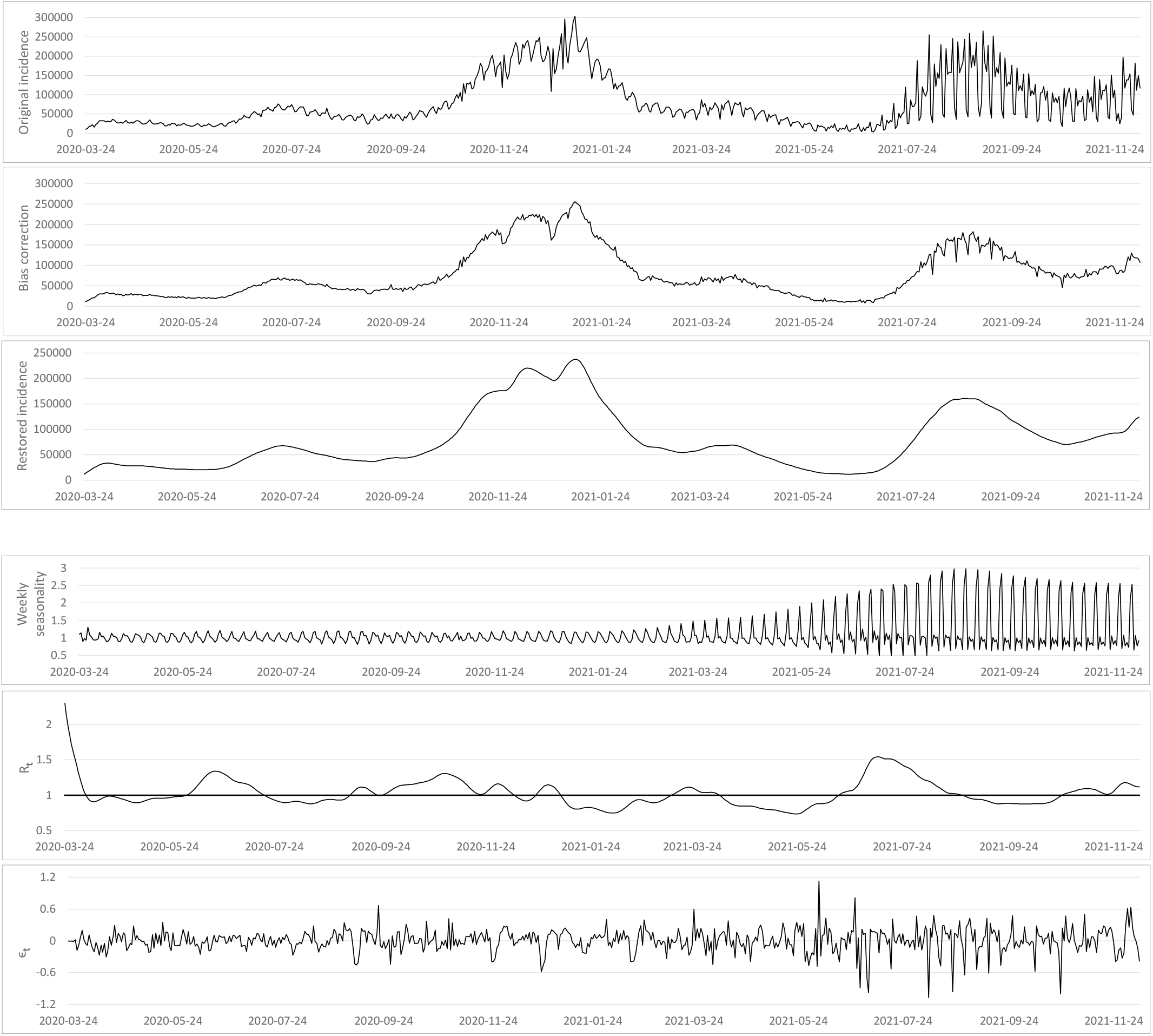
From top to bottom : (i) the original incidence curve 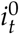 of USA, (ii) the incidence curve after bias correction *î*_*t*_, (iii) the restored incidence curve using the renewal equation 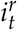, (iv) the weekly bias correction factors *q*_*t*_, (v) the reproduction number estimation *R*_*t*_ and (vi) the normalized error 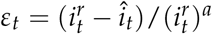, where *a* is the optimal exponent obtained by regression (see table A2).

**Figure A3.**
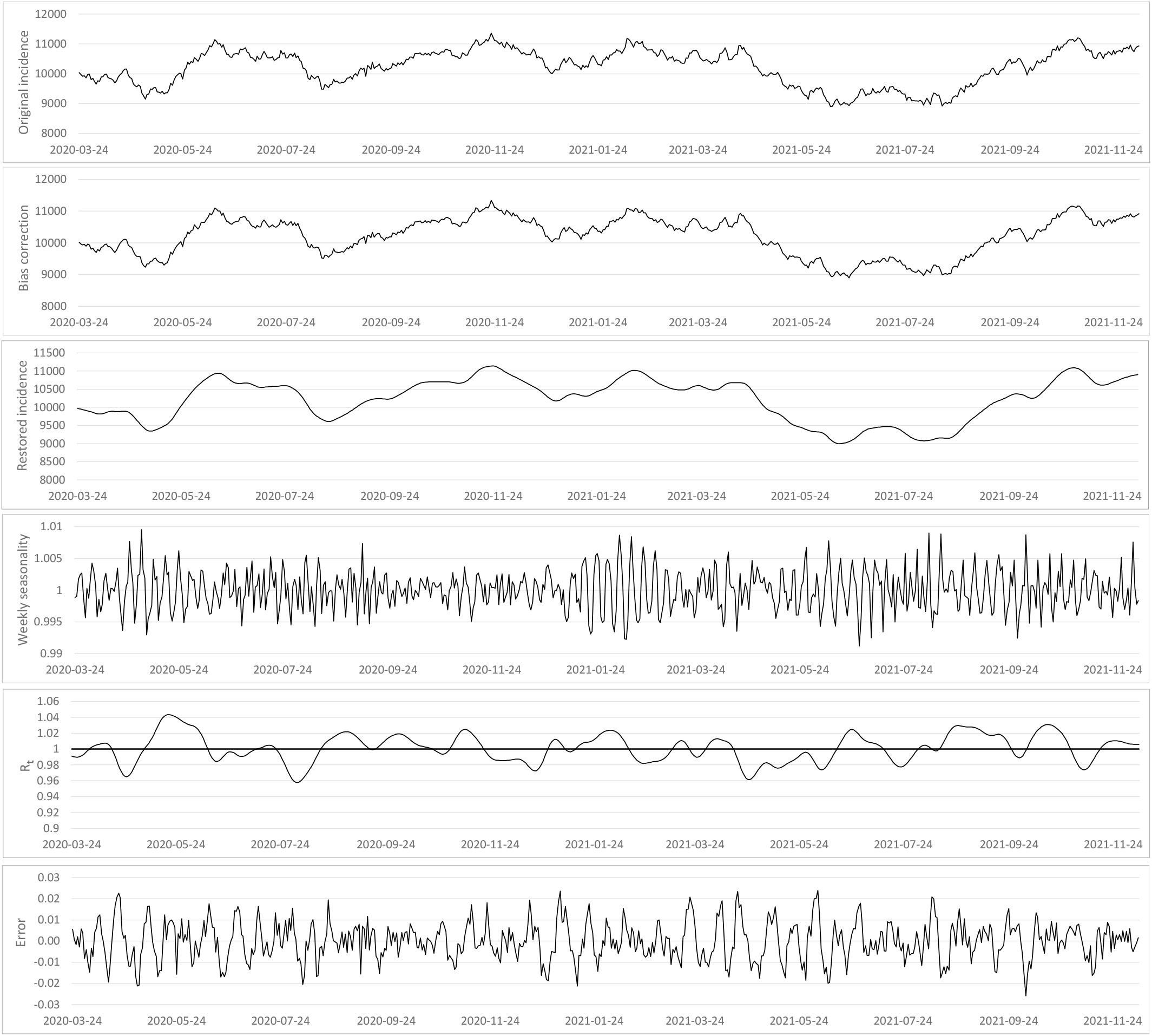
Control test, from top to bottom : (i) the test incidence curve 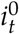 which is a Brownian motion, (ii) the test curve after bias correction *î*_*t*_, (iii) the restored incidence curve using the renewal equation 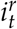, (iv) the weekly bias correction factors *q*_*t*_, (v) the reproduction number estimation *R*_*t*_ and (vi) the relative error 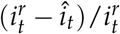. Both *R*_*t*_ and the weekly seasonality correction coefficients stay very close to 1, with means 1.001 and 1.00002, and standard deviations 1.7% and 0.3% respectively.

**Table A1.** Table with the mean and standard deviation of *ε*_*t*_ and the parameters of the best fit to the exponential distributions for 36 countries. The data of starred countries in the first three rows have undergone the festive bias correction.

| Country | Mean | Std | Location | Scale | Shape ($\beta$) |
| --- | --- | --- | --- | --- | --- |
|  |  |  | *Exponential* | *Exponential* | *Exponential* |
| **FRA\*** | -0.0283 | 0.8290 | -0.0286 | 0.5394 | 1.0000 |
| **DEU\*** | -0.0178 | 0.4785 | -0.0135 | 0.3433 | 1.0144 |
| **USA\*** | -0.0044 | 0.2169 | -0.0059 | 0.1537 | 1.0000 |
| FRA | 0.0109 | 1.0024 | -0.0316 | 0.6026 | 1.0000 |
| DEU | 0.0091 | 0.5143 | 0.0050 | 0.3458 | 1.0000 |
| USA | 0.0032 | 0.4779 | -0.0097 | 0.3003 | 1.0000 |
| ARG | 0.0025 | 0.4430 | -0.0286 | 0.3153 | 1.0000 |
| AUT | 0.0419 | 1.1030 | -0.0041 | 0.9035 | 1.2701 |
| BEL | 0.0413 | 1.2175 | -0.0366 | 0.8304 | 1.0000 |
| BRA | -0.0018 | 0.4825 | -0.0368 | 0.3312 | 1.0000 |
| CAN | 0.0068 | 1.2720 | -0.0252 | 0.8290 | 1.0000 |
| CHL | 0.0019 | 0.2960 | -0.0082 | 0.2138 | 1.0252 |
| COL | -0.0026 | 0.2006 | -0.0107 | 0.1490 | 1.0751 |
| CZE | 0.0116 | 0.5671 | -0.0415 | 0.3755 | 1.0000 |
| DNK | 0.0278 | 1.2446 | -0.0298 | 0.8126 | 1.0000 |
| GRC | 0.0218 | 1.2764 | -0.0410 | 0.8847 | 1.0000 |
| HUN | 0.0069 | 0.6600 | -0.0267 | 0.4410 | 1.0000 |
| IND | 0.0419 | 0.9891 | -0.0084 | 0.6786 | 1.0000 |
| IDN | -0.0015 | 0.3374 | -0.0140 | 0.2607 | 1.1466 |
| IRL | 0.0030 | 1.1778 | -0.0748 | 0.8252 | 1.0000 |
| ITA | 0.0368 | 1.1441 | 0.0141 | 0.7130 | 1.0000 |
| JPN | 0.0243 | 0.6647 | -0.0254 | 0.4515 | 1.0000 |
| MEX | -0.0318 | 1.7329 | -0.0955 | 1.1091 | 1.0000 |
| NPL | 0.0035 | 0.8994 | 0.0005 | 0.5652 | 1.0000 |
| NLD | 0.0437 | 0.7185 | -0.0404 | 0.4910 | 1.0000 |
| PHL | -0.0196 | 2.0401 | -0.0930 | 1.4011 | 1.0000 |
| POL | -0.0017 | 0.1911 | -0.0043 | 0.1268 | 1.0000 |
| ROU | 0.0063 | 0.9465 | -0.0011 | 0.5798 | 1.0000 |
| RUS | 0.0107 | 0.3383 | 0.0066 | 0.2270 | 1.0000 |
| SRB | 0.0675 | 1.0140 | 0.0758 | 0.7932 | 1.1728 |
| SVK | 0.0024 | 1.3671 | -0.0778 | 0.8194 | 1.0000 |
| ZAF | 0.0139 | 0.9110 | -0.0320 | 0.7059 | 1.1497 |
| ESP | 0.0637 | 1.6068 | -0.0047 | 1.0840 | 1.0000 |
| CHE | 0.0528 | 1.2228 | 0.0017 | 0.8667 | 1.0000 |
| THA | 0.0299 | 1.3738 | -0.0312 | 0.9374 | 1.0000 |
| TUN | 0.0123 | 1.3033 | -0.0845 | 0.9224 | 1.0000 |
| UKR | 0.0034 | 0.4117 | -0.0215 | 0.2586 | 1.0000 |
| ARE | 0.0108 | 0.4192 | -0.0127 | 0.3265 | 1.1588 |
| GBR | 0.0085 | 0.3304 | -0.0171 | 0.2163 | 1.0000 |

**Table A2.** Coefficients *a* and *b* for 38 countries of the log-log linear regression *ax* + *b* between restored incidence 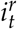 and the residual 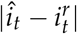 as displayed in Fig. 4. The Pearson correlation p-values given by the *stats* R package confirm a linear relation. The exponent *a* varies between 0.7 and 0.9. Stars* indicate countries with festive correction. The pvalues are slightly better with festive correction than without. The last row shows the results on the control curve, simulated as a Brownian process. Its large p-value discards a linear log-log relation, and the estimated values of *a* and *b* also stand far away from the estimated values for real incidence curves.

| Country | a | b | p-value | Country | a | b | p-value |
| --- | --- | --- | --- | --- | --- | --- | --- |
| **FRA\*** | 0.8074272 | -1.164141 | 2.01E-75 | FRA | 0.8136197 | -1.1710322 | 2.76E-71 |
| **DEU\*** | 0.8233846 | -1.496739 | 5.99E-95 | DEU | 0.8235076 | -1.5057318 | 3.01E-92 |
| **USA\*** | 0.9076139 | -2.264255 | 3.16E-42 | USA | 0.8638492 | -1.7287377 | 6.37E-37 |
| ARG | 0.8340299 | -1.5574878 | 1.71E-101 | AUT | 0.6628437 | -0.5661912 | 3.45E-86 |
| BGD | 0.9104934 | -2.5672893 | 6.14E-56 | BEL | 0.7184413 | -0.6589731 | 3.65E-61 |
| BRA | 0.8906214 | -1.536314 | 1.03E-58 | CAN | 0.7240632 | -0.6726824 | 2.96E-44 |
| CHL | 0.8349688 | -1.9543089 | 2.64E-40 | COL | 0.9175985 | -2.2638884 | 3.03E-112 |
| CZE | 0.8520268 | -1.4708978 | 2.88E-133 | DNK | 0.6900743 | -0.6284769 | 2.78E-68 |
| GRC | 0.6555842 | -0.5683038 | 2.58E-102 | HUN | 0.7838904 | -1.3618843 | 4.47E-142 |
| IND | 0.7042499 | -0.8457334 | 8.20E-68 | IDN | 0.8406915 | -1.7674138 | 5.30E-97 |
| IRL | 0.7043354 | -0.5484242 | 1.35E-89 | ITA | 0.6964125 | -0.8659193 | 2.53E-71 |
| JPN | 0.7222903 | -1.2445353 | 5.65E-85 | MEX | 0.725394 | -0.4661005 | 1.76E-32 |
| NPL | 0.7548857 | -1.0559482 | 1.42E-55 | NLD | 0.7494921 | -1.1280471 | 3.35E-96 |
| PHL | 0.6715338 | -0.1103984 | 1.90E-47 | POL | 0.9306078 | -2.6041615 | 3.02E-133 |
| ROU | 0.6920366 | -1.0282145 | 4.11E-77 | RUS | 0.7212814 | -2.0048746 | 4.05E-26 |
| SRB | 0.628712 | -0.65103 | 6.76E-92 | SVK | 0.7381511 | -0.7853881 | 8.53E-164 |
| ZAF | 0.7275793 | -0.7811203 | 9.48E-69 | ESP | 0.6806819 | -0.3916179 | 2.03E-42 |
| CHE | 0.6138378 | -0.5491828 | 1.38E-75 | THA | 0.7110685 | -0.4672682 | 1.63E-222 |
| TUN | 0.7539949 | -0.503523 | 3.03E-163 | TUR | 0.8998264 | -2.658924 | 1.32E-68 |
| UKR | 0.8172308 | -1.8996555 | 1.75E-70 | ARE | 0.7511088 | -1.5460453 | 3.80E-52 |
| GBR | 0.8705096 | -1.9395546 | 1.70E-96 | **World** | 0.7631129 | -1.1389749 | 0.0000000 |
| **Brownian** | -1.0155743 | 13.3969412 | 0.0844 |  |  |  |  |

**Figure A4.**
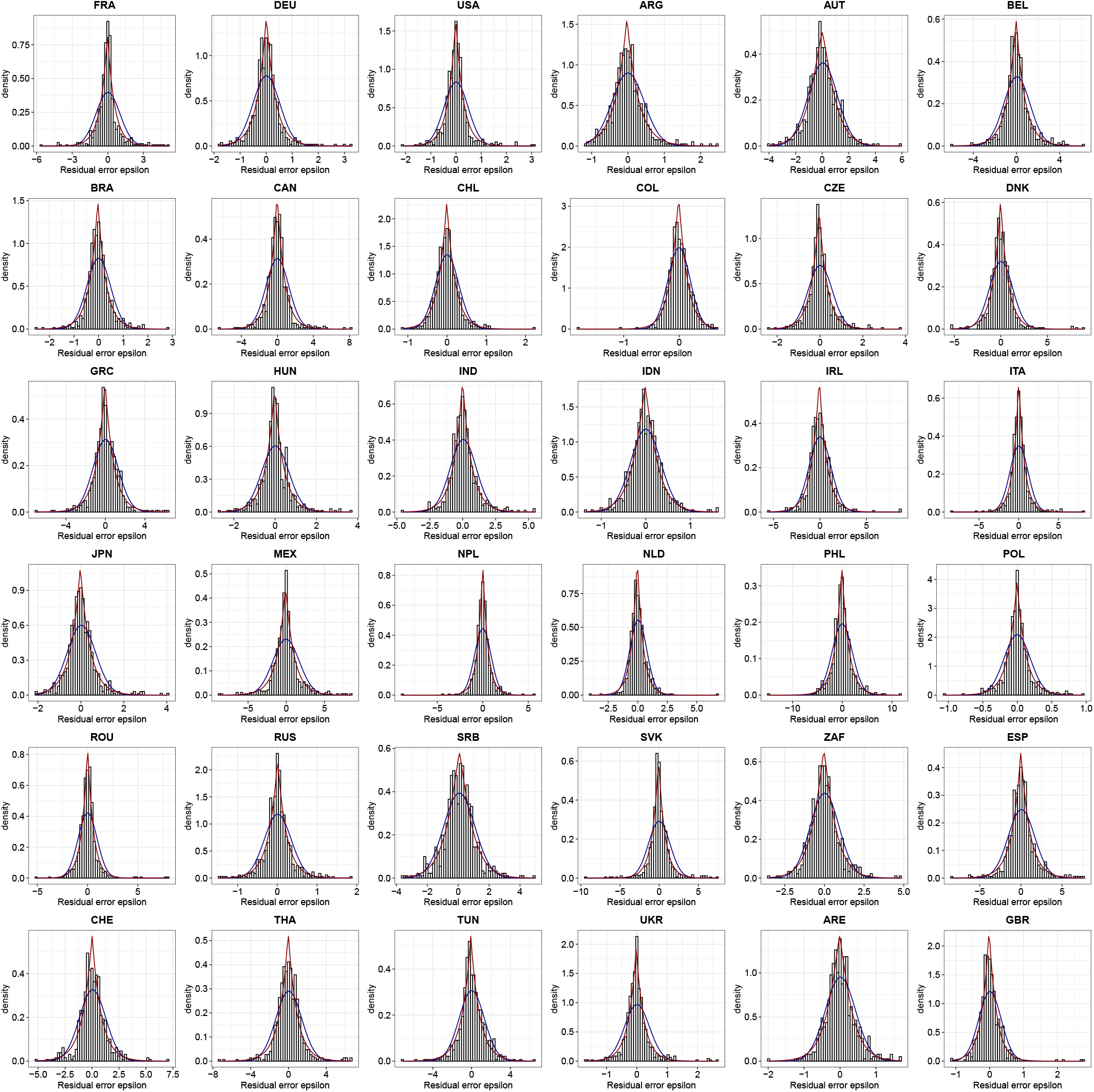
Fit of exponential distributions for 36 countries. In red, the best fitting exponential distribution with shape larger or equal to 1. In black, the best fitting normal law.

**Figure A5.**
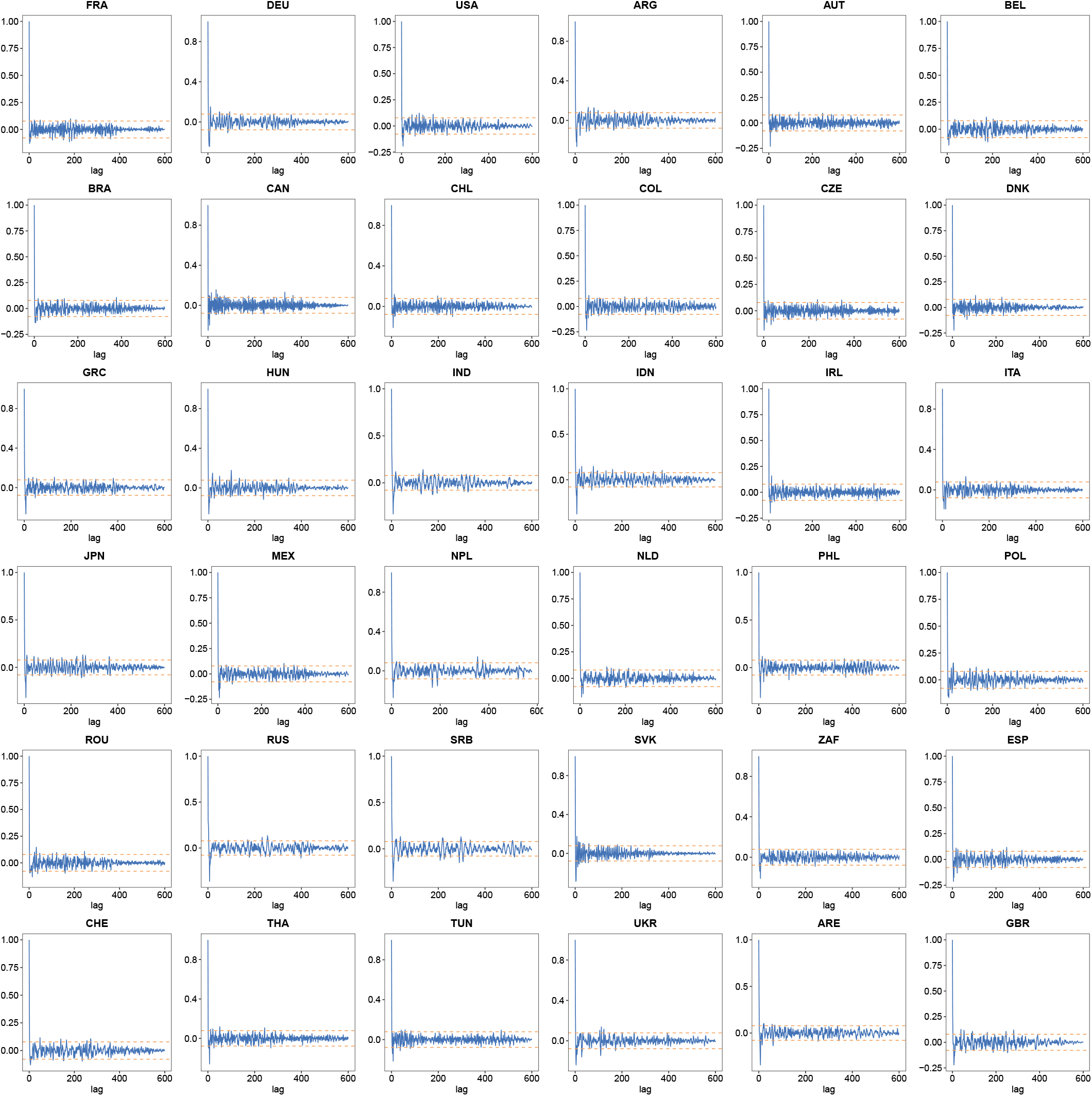
Autocorrelation of the normalized error *ε*_*t*_ using the R-software functionalities (acf() function) for 36 countries. The dotted lines give the 95% confidence interval for non-correlation.

**Figure A6.**
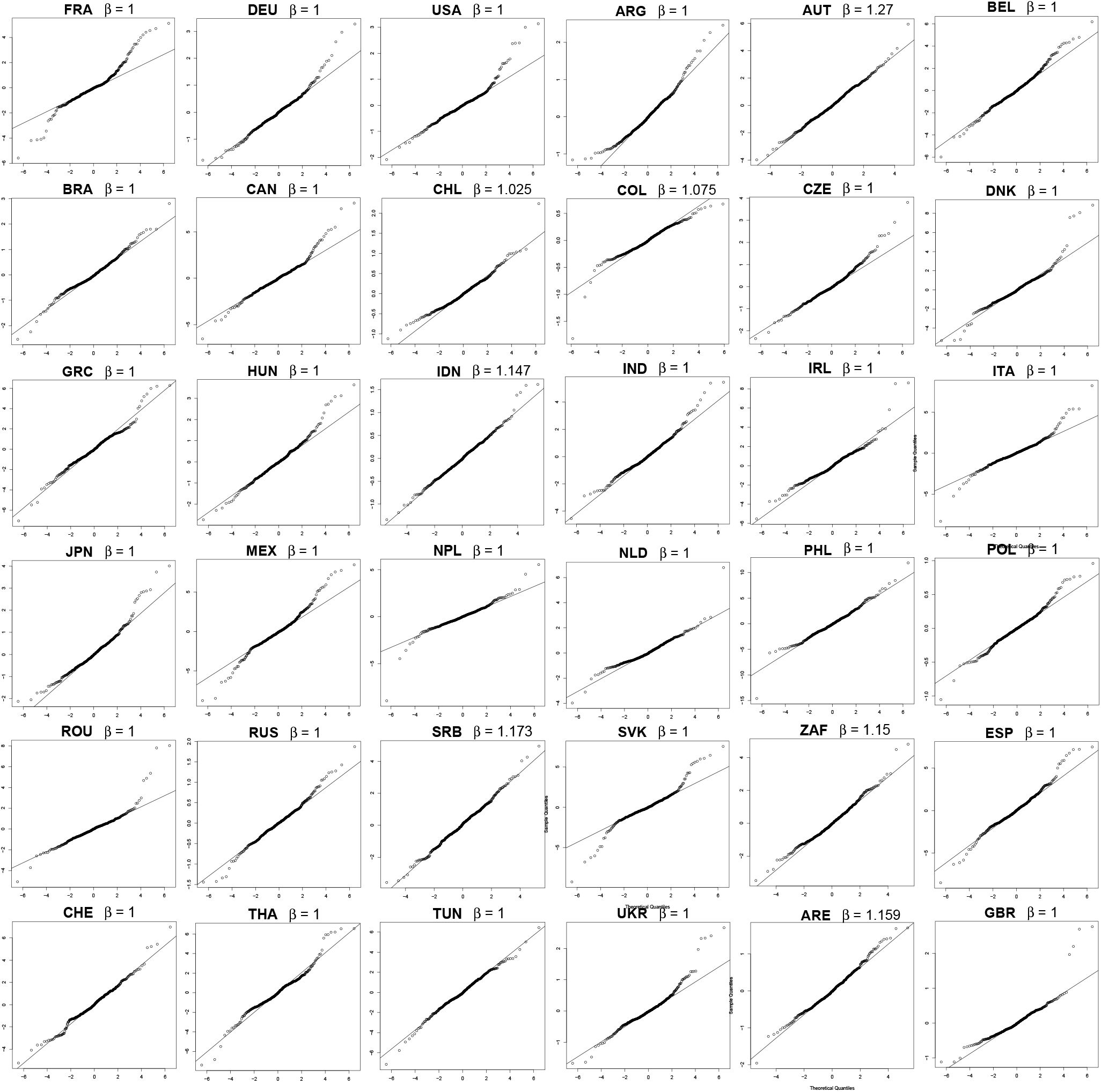
Quantile-quantile plot with 36 countries comparing *ε*_*t*_ (without using the festive day correction) with the optimal exponential distribution using the R-package *normalp*. In the horizontal axis we show the theoretical quantiles and in the vertical axis, the sample quantiles. Note that the exponential distribution shape parameter *β*, indicated on the graphs can have values >1.

## Notes

### Competing Interest Statement

The authors have declared no competing interest.

### Funding Statement

This study did not receive any funding

### Summary of Updates

We change the title and we add a detailed review of related papers.

